# Characteristics of anti-SARS-CoV-2 antibodies in recovered COVID-19 subjects

**DOI:** 10.1101/2020.09.11.20192690

**Authors:** Angela Huynh, Donald M. Arnold, James W. Smith, Jane C. Moore, Ali Zhang, Zain Chagla, Bart J. Harvey, Hannah D. Stacey, Jann C. Ang, Rumi Clare, Nikola Ivetic, Vasudhevan T. Chetty, Dawn ME Bowdish, Matthew S. Miller, John G. Kelton, Ishac Nazy

## Abstract

Coronavirus Disease 2019 (COVID-19) is a global pandemic caused by the novel severe acute respiratory syndrome coronavirus 2 (SARS-CoV-2). While detection of SARS-CoV-2 by polymerase chain reaction with reverse transcription (RT-PCR) is currently used to diagnose acute COVID-19 infection, serological assays are needed to study the humoral immune response to SARS-CoV-2. SARS-CoV-2 IgG/A/M antibodies against SARS-CoV-2 spike (S) protein and its receptor-binding domain (RBD) were characterized using an enzyme-linked immunosorbent assay (ELISA) and assessed for their ability to neutralize live SARS-CoV-2 virus in recovered subjects who were RT-PCR-positive (n=153), RT-PCR-negative (n=55), and control samples collected pre-COVID-19 (n=520). Anti-SARS-CoV-2 antibodies were detected in 90.9% of resolved subjects up to 180 days post-symptom onset. Anti-S protein and anti-RBD IgG titers correlated (r= 0.5157 and r = 0.6010, respectively) with viral neutralization. Of the RT-PCR-positive subjects, 22 (14.3%) did not have anti-SARS-CoV-2 antibodies; and of those, 17 had RT-PCR cycle threshold (Ct) values >27, raising the possibility that these indeterminate results are from individuals who were not infected, or had mild infection that failed to elicit an antibody response. This study highlights the importance of serological surveys to determine population-level immunity based on infection numbers as determined by RT-PCR.

## Introduction

Coronavirus disease 2019 (COVID-19) is caused by the novel severe acute respiratory syndrome coronavirus 2 (SARS-CoV-2) [1]. Asymptomatic and pre-symptomatic virus transmission is one of the biggest challenges with the global pandemic [2]. It is estimated up to 80% of people infected with COVID-19 have none or mild symptoms and asymptomatic transmission accounts for half of all COVID-19 infections [3-5]. Approximately 20% of symptomatic infections are severe, disproportionately impacting the elderly and patients with underlying health conditions [6, 7].

Laboratory diagnosis of COVID-19 is made using polymerase chain reaction with reverse transcription (RT-PCR) to detect viral mRNA from nasal or throat swabs [8-11]. Viral RNA is detectable as early as the first day of symptom onset and peaks within the first week of symptom onset [9]. The SARS-CoV-2 spike (S) protein, specifically the receptor-binding domain (RBD), facilitates viral entry into the cell via the angiotensin-converting enzyme-2 (ACE2) receptor on host cells [12-15]. Most people with a confirmed RT-PCR diagnosis of SARS-CoV-2 infection develop IgG, IgA and IgM antibodies against S protein within 1–2 weeks of symptom onset and continue to circulate after initial viral clearance [16-21].

Serological studies have shown IgG antibodies to SARS-CoV-2 S protein and RBD are detected in the circulation of greater than 90% of infected subjects by 10–11 days post-symptom onset [16, 18, 20, 22]. Virus-specific neutralizing antibodies, either induced through infection or vaccination, can block viral infection [23]. Although antibodies may be generated against multiple domains within the S protein, most neutralizing antibodies and highly potent monoclonal antibodies target the RBD [15, 24]. In this report, we profile the IgG, IgA, and IgM responses to the SARS-CoV-2 S protein and RBD in a cross-sectional serological study involving resolved COVID-19 infection. We also compared antibody levels with viral neutralization and RT-PCR results.

## Methods and Materials

### Study Design

Subjects who recovered from COVID-19 infection were identified by treating physicians, public health officials and through hospital discharge databases that included hospitals in Hamilton, Ontario, Canada (Hamilton General Hospital, Juravinski Hospital, McMaster University Medical Centre, and St. Joseph’s Healthcare). The study inclusion criteria were ≥18 years of age, either testing positive or negative for COVID-19 in the RT-PCR, with no exclusion criteria.

Participants with RT-PCR-positive results for SARS-CoV-2 and had since recovered (resolved RT-PCR-positive, n=153), and subjects who experienced symptoms but tested negative by RT-PCR (RT-PCR-negative, n=55) were included. Participants were interviewed by phone and self-reported their age, sex, symptom onset date and RT-PCR test date and result. Pre-COVID-19 control samples were selected from healthy donors (n=37) and samples sent for the ITP Registry biobank from the McMaster Platelet Immunology Laboratory drawn prior to November 2019 (pre-COVID-19, n=483) before documented local community transmission of SARS-CoV-2. Serum was collected by venipuncture and cryopreserved until use. This study was approved by the Hamilton Integrated Research Ethics Board (HIREB) and written informed consent was obtained from all participants.

### Production of recombinant SARS-CoV-2 S protein and RBD

A detailed protocol outlining protein production can be found in a study by Stadlbauer *et al* [25] and is outlined in the Supplemental methods.

### Measuring SARS-CoV-2 antibodies

Microtitre well plates (384 wells, Nunc Maxisorp, Rochester, NY, USA) were coated with 25μL/well of S protein (5μg/mL) or RBD (2μg/mL) suspended in 50 mM carbonate-bicarbonate buffer (pH 9.6). The plates were then blocked with 100µL/well of 3% skim milk prepared in phosphate buffered saline (PBS) with 0.05% Tween 20 at room temperature for 2-hours. The blocking solution was removed, and diluted serum samples (1/100 prepared in 1% skim milk in PBS/0.05% Tween 20) in technical duplicates were added to the plates and incubated for 1-hour at room temperature. The plates were washed twice with PBS/0.05% Tween 20 and thrice with PBS. Bound human antibodies (IgG, IgA, or IgM) were detected with 25μL/well of alkaline phosphatase conjugated goat anti-human IgG (γ-chain-specific, 1/2000, Jackson ImmunoResearch Laboratories, Inc, Westgrove, PA, USA), goat anti-human IgA (α-chain-specific; 1/500, Jackson ImmunoResearch Laboratories, Inc, Westgrove, PA, USA) antibody, or goat anti-human IgM (μ-chain-specific; 1/1000, Jackson ImmunoResearch Laboratories, Inc, Westgrove, PA, USA) antibody prepared in PBS/0.05% Tween 20 with 1% skim milk. Plates were washed as before and followed with the addition of 50µL substrate (4-nitrophenylphosphate disodium salt hexahydrate in diethanolamine (MilliporeSigma, St. Louis, MO, USA). The optical density (OD) was read at 405nm and 490nm (as a reference) measured using a BioTek 800TS microplate reader (BioTek, Winooski, VT, USA). The assay cut-off was determined as the mean and 3 standard deviations (SD) of the pre-COVID-19 control population. Data are shown as a ratio of observed OD to the determined assay cut-off OD. OD ratio values above 1 ratio were considered positive in the SARS-CoV-2 ELISA. Results for optimization of antigens, serum concentrations for the ELISA and its comparisons to commercially available assays can be found in the Supplemental data.

### PCR Cycle threshold (Ct) values of resolved COVID-19 samples

RT-PCR Ct values were retrieved from a subset of resolved subjects’ original test date RT-PCR (n=54). A detailed protocol of the in-house RT-PCR run by the Hamilton Regional Laboratory Medicine Program virology lab is outlined in the Supplemental methods.

### Detecting neutralizing antibodies for SARS-CoV-2

Vero E6 cells (ATCC CRL-1586) were seeded at a density of 2.5×10^4^ cells per well in opaque 96 well flat-bottom plates (Costar) in complete DMEM (supplemented with 10% FBS, 1% L-glutamine, 100U/ml penicillin-streptomycin). After 24-hours of incubation, serum (resolved and RT-PCR-negative subjects) was inactivated by incubating at 56°C for 30 minutes, then diluted 1:10 in low serum DMEM (supplemented with 2% FBS, 1% L-glutamine, 100U/mL penicillin-streptomycin), followed by a 1:2 dilution series in 96 well U-bottom plates resulting in a final volume of 55μL diluted serum per well. An equal volume of SARS-CoV-2/SB3-TYAGNC consisting of 330 PFU per well was then added to the diluted serum and the serum-virus mixture was incubated at 37°C for 1-hour. Next, the Vero E6 culture media was then replaced with 100 μL of the serum-virus mixture and was incubated at 37°C for 72-hours. The plates were read by removing 50μL of culture supernatant and adding 50μL of CellTiter-Glo 2.0 Reagent (Promega, G9243) to each well. The plates were then shaken at 282cpm at 3 mm diameter for 2-minutes, incubated for 5-minutes at room temperature and luminescence was read using a BioTek Synergy H1 microplate reader with a gain of 135 and an integration time of 1-second. Results are expressed as geometric microneutralization titers at 50% (MNT50).

### Statistical Analyses

Descriptive statistics were used to summarize the IgG, IgA, and IgM binding to S protein and RBD as measured by mean OD across antigen and technical replicates. Differences between data were tested for statistical significance using the paired or unpaired t-test and the Mann-Whitney test. P-values are reported as 2-tailed. Correlations were calculated using standard Pearson correlation. All statistical analyses were conducted using GraphPad Prism (version 7.0a, GraphPad Software, San Diego, USA).

## Results

### Study Demographics

Resolved (RT-PCR-positive, n=153) samples were collected between 7-211 days post-symptom onset. Median age of the resolved RT-PCR-positive subjects was 49 years (range: 18 – 82) and 95 subjects (62.1%) were female. COVID-19 negative subjects (RT-PCR-negative, n=55) samples were collected between 7-246 days post-symptoms (Table 1). The median age of the RT-PCR-negative subjects was 49 years (range: 20 – 89) and 39 (70.9%) were female (Table 1). Eleven (7.2%) of the RT-PCR-positive resolved subjects and 37 (67.3%) of the RT-PCR-negative subjects were asymptomatic before testing.

**Table 1:** Clinical Characteristics of Study Participants.

|  | Pre-COVID-19<br>controls<br>(n = 520) | Resolved<br>COVID-19<br>subjects<br>(n = 153) | RT-PCR-negative<br>subjects<br>(n = 55) |
| --- | --- | --- | --- |
| Ages (years) | - | 18 to 82<br>(median=49) | 20 to 89<br>(median=49) |
| Gender |  |  |  |
| Male (%) | - | 58 (37.9) | 16 (29.1) |
| Female (%) | - | 95 (62.1) | 39 (70.9) |
| Hospitalization Status |  |  |  |
| Never hospitalized (%) | - | 145 (94.8) | 55 (100) |
| Hospitalized (%) | - | 8 (5.2) | 0 |
| Sample Collection Dates | - | May 2020 – November 2020 |  |
| SARS-CoV-2 RT-PCR positivity |  |  |  |
| Positive | - | 153 | 0 |
| Negative | - | 0 | 55 |
| Presence of symptoms |  |  |  |
| Symptomatic (%) | - | 142 (92.8) | 37 (67.3) |
| Asymptomatic (%) | - | 11 (7.2) | 18 (32.7) |
| **Days post-symptom onset at**<br>**collection (days)** | - | 19 – 227<br>(median=130.5) | 7 – 260<br>(median=141.5) |

### Detecting SARS-CoV-2 antibodies in resolved COVID-19 subjects

To study the antibody response to SARS-CoV-2, we tested for IgG, IgA, and IgM antibodies to the S protein and RBD in pre-COVID-19 controls (n=520), resolved COVID-19 subjects (n=153), and RT-PCR-negative subjects (n=55). Pre-COVID-19 controls (n=520) were used to determine the background reactivity to the S protein and RBD using samples from individuals drawn before COVID-19 (pre-November 2019). The cut-off was determined as the mean plus 3SD of the OD readings in the pre-COVID-19 control population. Most pre-COVID-19 controls had only background reactivity for both the full-length S protein and RBD (IgG = 98.9%, n= 514/520 and 98.5%, n = 512/520 below established cut-off, respectively). Each antigen and antibody class had a few pre-COVID-19 controls that tested positive for the antibodies based on the determined cut-off. The majority of pre-COVID-19 controls that tested positive within the groups had IgM against both S protein and RBD, 1.5% (n = 8/520 testing antibody-positive) and 2.1% (n= 11/520 antibody-positive), respectively. Some control samples were positive in the S protein- and RBD-specific IgA assays: 1.3% (n = 7/520 antibody-positive) and 1.0% (n = 5/520 antibody-positive), respectively. Antigen concentration of S protein and RBD and serum dilutions were optimized by testing known COVID-19-positive and pre-COVID-19 samples (see Supplemental data).

Of the 153 resolved COVID-19 subjects tested, 131 (85.6%) tested positive for antibodies against SARS-CoV-2 (IgG, IgA, or IgM antibodies against the S protein or RBD, Table 2) and 22 (14.4%) did not have detectable antibodies against SARS-CoV-2. Of the 55 RT-PCR-negative subjects, three had reactivity to the SARS-CoV-2 antigens (5.5%, Table 2). Most resolved subjects tested positive for anti-S protein and anti-RBD IgG (n=130/153 testing positive (85.0%) and n=119/153 testing positive (77.8%), respectively, Supplemental data). In addition, some of the same resolved subjects also tested positive for anti-S protein IgA (60.1%, 92/153 antibody-positive), anti-S protein IgM (35.3%, 54/153 antibody-positive), and anti-RBD IgA (24.2%, 37/153 antibody-positive) and IgM (19.6%, 30/153 antibody-positive).

**Table 2:** Cross-sectional analysis of RT-PCR and SARS-CoV-2 Antibody Testing.

|  | **RT-PCR-positive** | **RT-PCR-negative** |
| --- | --- | --- |
|  | **(Resolved, n=153)** | **(n=55)** |
| **SARS-CoV-2** | 131/153 | 3/55 |
| **Antibody positive (%)** | 85.6 | 5.5 |
| **SARS-CoV-2** | 22/153 | 52/55 |
| **Antibody negative (%)** | 14.4 | 94.5 |

### High Ct counts found in resolved subjects who were SARS-CoV-2 antibody-negative

To further understand RT-PCR-positive COVID-19 subjects that tested negative for anti-SARS-CoV-2 antibodies, 22/153 (14.4%) resolved subjects were further investigated. The Ct values from the initial RT-PCR test were obtained for 54/153 (35.3%) of the resolved participants, including 18/22 (81.8%) who were RT-PCR-positive but antibody-negative. The Ct values for the RT-PCR-positive/antibody-negative subjects (n=18) ranged from 16.00 to 37.38, with a mean of 32.29 ± 4.647, whereas the mean of the subjects who were RT-PCR-positive/antibody-positive was 22.92 ± 5.177 (range = 14.99 – 34.94, n=36, Figure 1A, 1B). Sera from 13 of 18 resolved subjects who tested RT-PCR-positive/antibody-negative were collected within 60 days after initial RT-PCR test, within the reported optimal time for anti-SARS-CoV-2 antibodies (Figure 1C) [18, 20, 21, 26].

**Figure 1:**
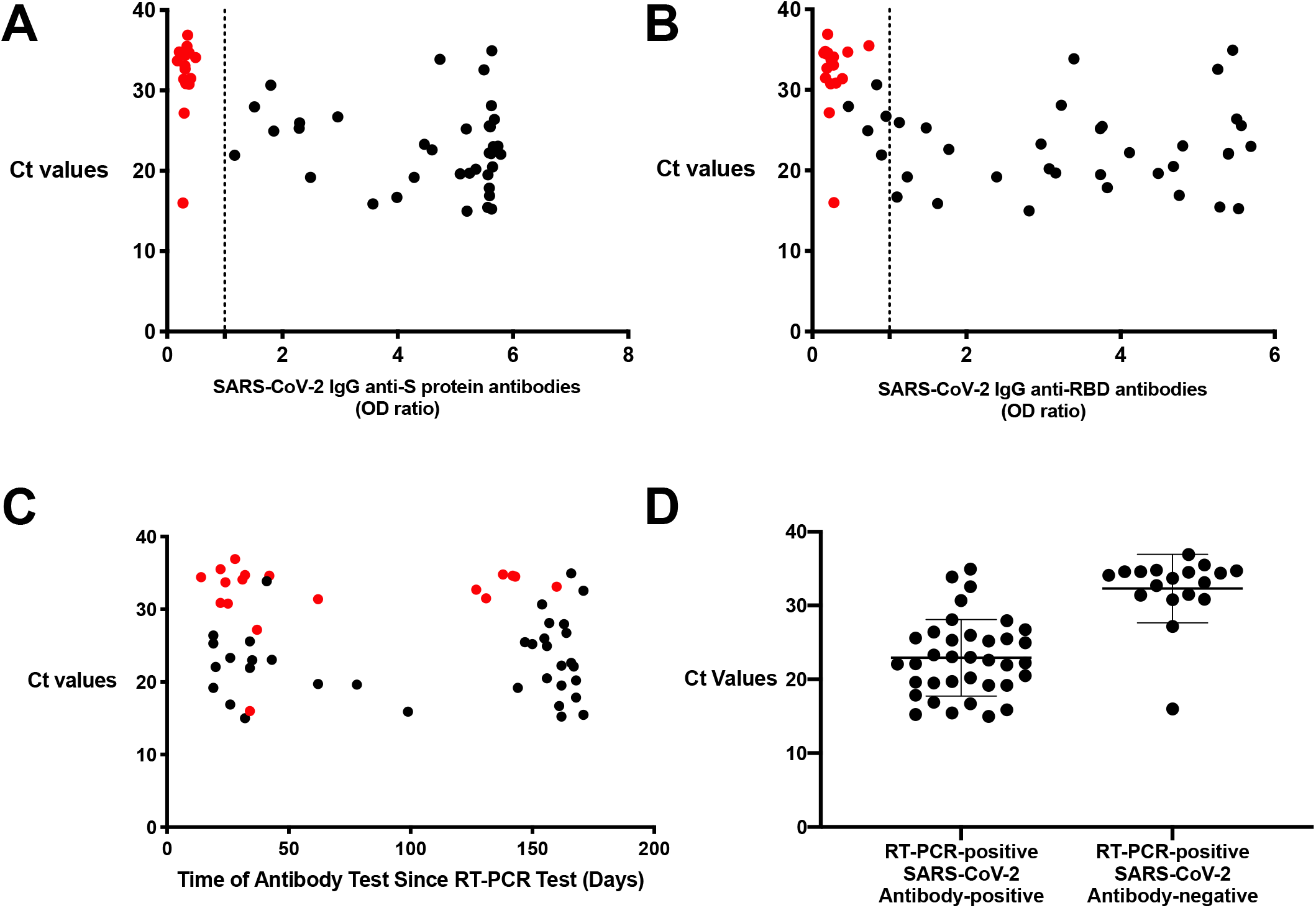
Ct values are variable in resolved subjects who test SARS-CoV-2 antibody negative or positive. Ct values of a subset of resolved subjects (n=54) were compared to their corresponding SARS-CoV-2 (A) anti-S protein IgG and (B) anti-RBD IgG. (C) Ct values were then compared to the subjects’ respective time since initial RT-PCR test. (D) Ct values for RT-PCR-positive in antibody-positive and antibody-negative were compared. Values are shown as a ratio of observed optical density to the determined assay cut-off optical density or time since RT-PCR test until blood donation compared to absolute Ct values. Red circles indicate resolved samples who were RT-PCR-positive/antibody-negative.

During assay validation, 14 resolved subjects (9 antibody-positive and 5 antibody-negative) were tested in the commercially available EUROIMMUN Anti-SARS-CoV-2 ELISA and the Ortho Clinical Diagnostics COVID-19 IgG Antibody Test. There was 100% correlation between the commercial reference assays and our in-house developed assays. Three of the 14 in validation testing were RT-PCR-positive/antibody-negative and had high Ct value samples and were confirmed to be negative for SARS-CoV-2 antibodies in the commercial reference assays (Supplemental data).

### Persistence of anti-S protein and anti-RBD antibodies in resolved subjects

IgG antibodies against S protein and RBD were detected in samples collected >120 days from symptom onset (Figure 2A)[5, 16, 18, 20]. Anti-S protein IgG was found in all resolved participants collected between 60 and 120 days (23/153 resolved subjects). In the resolved patients collected between 120 and 180 days from symptom onset, there was a decrease in the percentage of antibody-positive samples when compared to the previous time bin (55/60 samples, 91.7%). Of the resolved subjects tested >180 days of symptom onset (n=11), 90.9% had an anti-S protein IgG antibody. However, IgG levels against RBD demonstrated a 14.8% decrease in subjects who were antibody-positive between 120-180 days when compared to the groups before 120 days, and a 30% decrease in subjects with antibody after 180 days (Figure 2A). In contrast, IgA and IgM to both antigens were much less sustained (IgA - Figure 2B, IgM – Figure 2C). After reaching a maximum in the 0–60 days bin, there was a clear and continuous decline throughout the time series such that after 180 days, the anti-S protein and anti-RBD IgA levels in subject groups declined by 30% and 80% respectively, while IgM levels for both antigens declined by 90% (IgA - Figure 2B, IgM – Figure 2C).

**Figure 2:**
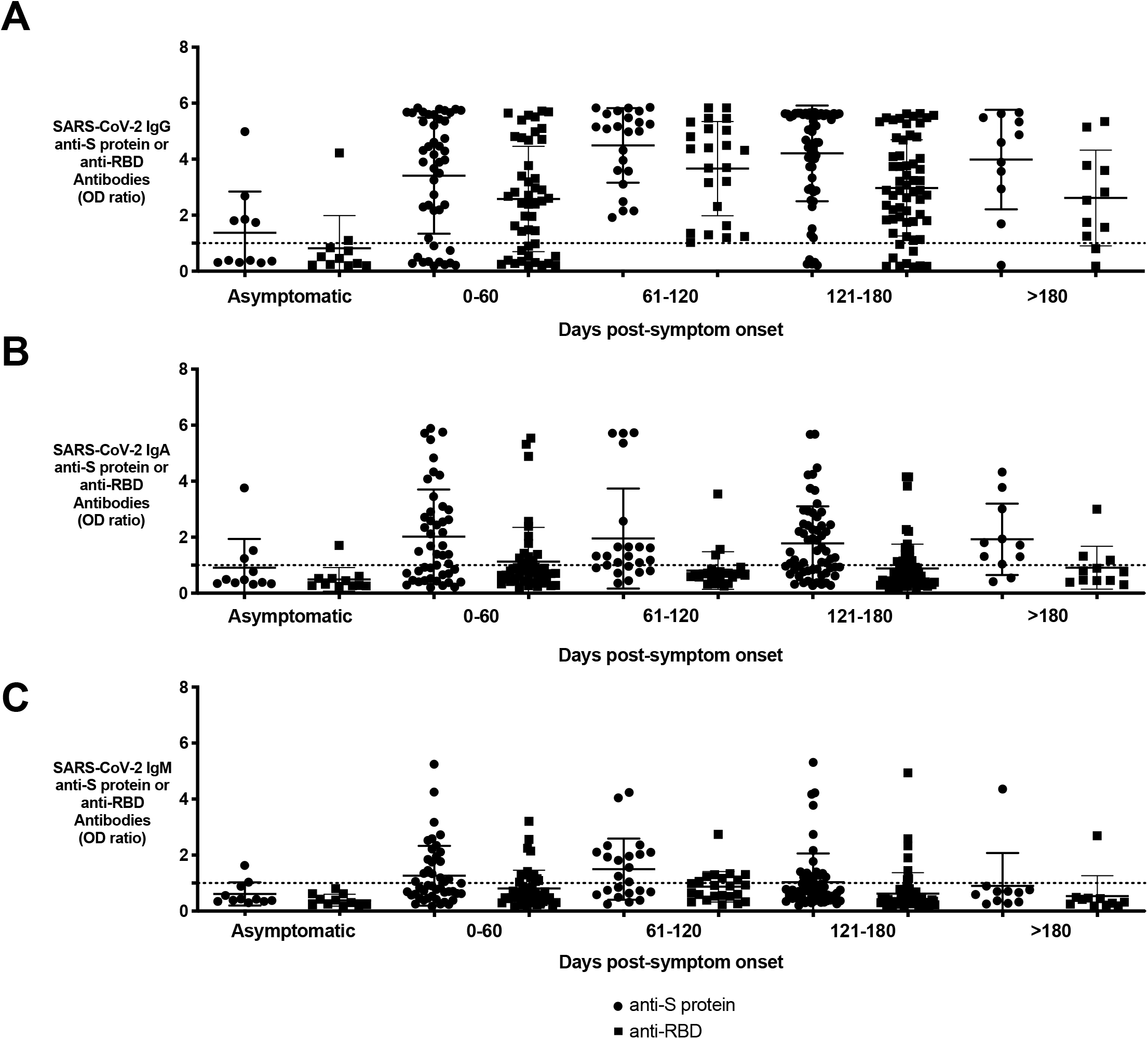
Persistence of SARS-CoV-2 antibodies in resolved subjects based on time of blood collection post-symptom onset. Resolved subjects (n=153) were grouped based on days post-symptom onset showing levels of (A) anti-S protein and anti-RBD IgG, (B) IgA, and (C) IgM displayed as dot plots. Days post-symptom onset are binned in 60-day increments and are compared to asymptomatic resolved subjects. Circles represent anti-S protein antibodies and squares represent anti-RBD antibodies. Anti-S protein IgG was found in all resolved participants collected between 60 and 120 days (23 of 153 total resolved subjects). In the resolved patients collected between 120 and 180 days from symptom onset, there was a decrease in the percentage of antibody-positive samples when compared to the previous time bin (55 of 60 samples, 91.7%).

### Investigation of neutralizing SARS-CoV-2 antibodies

Neutralization potency was measured using a microneutralization assay with live SARS-CoV-2 virus. In all resolved subjects, the presence of high titers of anti-S protein and anti-RBD IgG moderately correlated with higher titers of neutralizing antibodies (Figure 3A, 3B, r=0.5157, p<0.0001 anti-S protein IgG and r=0.601, p<0.0001 anti-RBD IgG). Weaker correlations were found between neutralizing antibody titers and anti-S protein IgA (r=0.4507), IgM (r=0.4443), and anti-RBD IgA (r=0.3055), IgM (r=0.3365). Geometric MNT50s ranged from below detection limit (MNT50 = 5) to MNT50=1280. Resolved subjects who were only antibody-positive for anti-S protein IgG but not antibody-positive for anti-RBD IgG antibodies (n=11) either had lower neutralizing antibody levels (mean MNT50 = 19.5, range 5 - 80) or were undetectable. No temporal trends were observed based on this cross-sectional study of resolved subjects, however neutralizing antibodies in resolved subjects were detected as far as 180 days post-symptom onset (Figure 3C). After 180 days, 3/11 (27.3%) of subjects tested for neutralizing antibodies had a level above MNT50=160.

**Figure 3:**
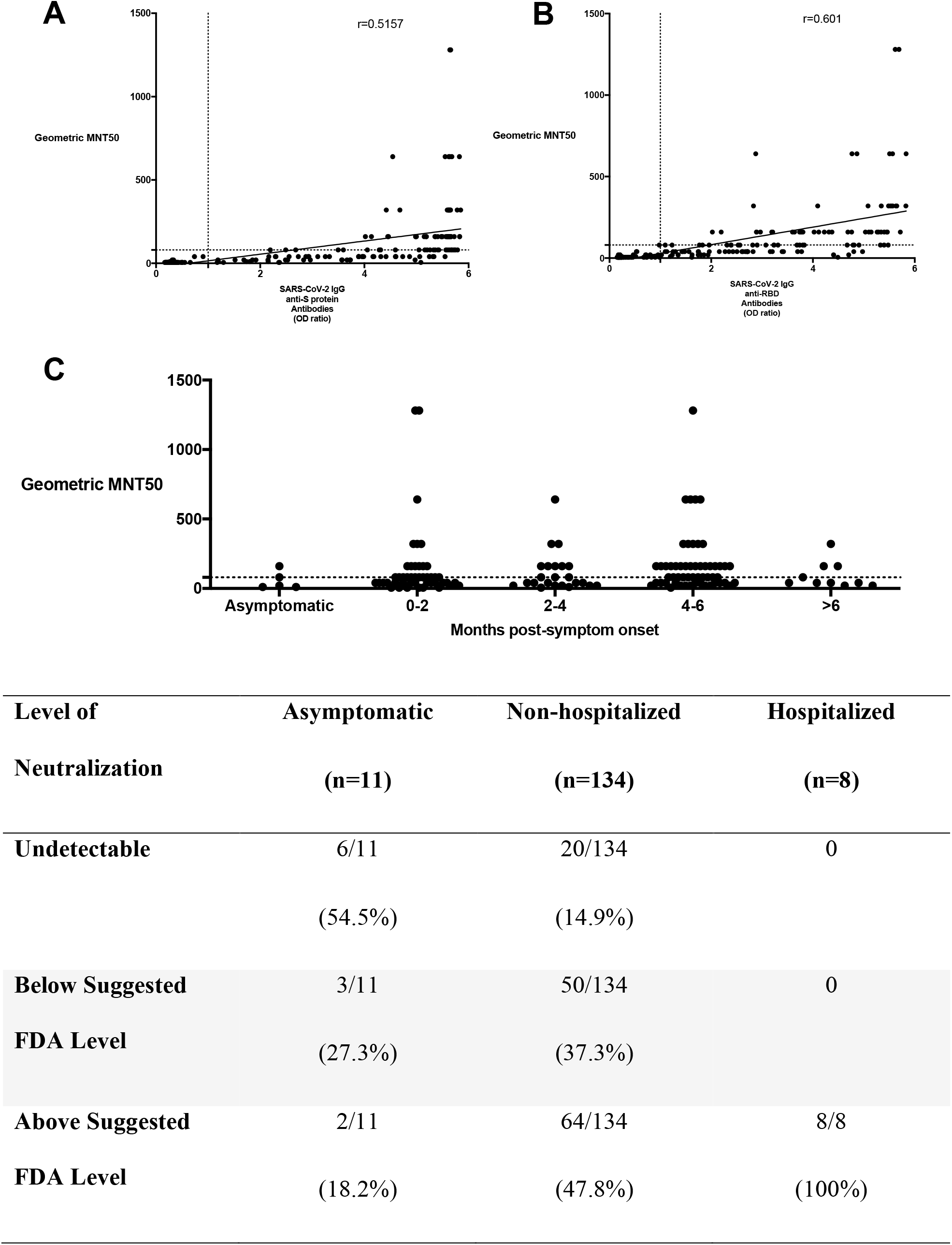
Neutralizing SARS-CoV-2 antibodies against S protein and RBD in IgG, IgA, and IgM found in variable levels in resolved and RT-PCR-negative study participants. Neutralizing SARS-CoV-2 antibody titers from resolved subjects (n=153) were measured in the microneutralization assay and compared to (A) anti-S protein and (B) anti-RBD IgG antibody levels as measured in the SARS-CoV-2 ELISA. Neutralizing antibody titers are expressed as geometric MNT50 values (y-axis). ELISA values are shown as a ratio of observed optical density to the determined assay cut-off optical density (x-axis). (C) Neutralizing SARS-CoV-2 antibody titers from resolved subjects (n=153) were measured in the microneutralization assay were compared to their days since symptom onset. Values above 1 ratio are considered positive in the SARS-CoV-2 ELISA.

### SARS-CoV-2 antibody profile of asymptomatic, non-hospitalized, and hospitalized resolved subjects

Of the 153 resolved subjects tested, eight (5.2%) were hospitalized for SARS-CoV-2 infection and were positive for anti-SARS-CoV-2 antibodies. All hospitalized subjects had detectable anti-RBD and anti-S protein IgG, and anti-S protein IgA antibodies in their serum (Figure 4A and 4B). The levels of anti-S protein and anti-RBD IgG (mean OD_405_ ratio) in resolved subjects who were hospitalized were significantly higher than the non-hospitalized resolved population (anti-S protein IgG; 5.667±0.065 vs. 3.836±1.831, p<0.001; anti-RBD IgG 5.240±0.8483 vs. 2.758±1.730, p<0.001). All other antigen and antibody classes were not significantly different between hospitalized and non-hospitalized resolved subjects. When compared to resolved hospitalized subjects (range: 3.209 – 5.831), there was a larger spread in antibody levels in the non-hospitalized population (range: 0.1508 – 5.828; Figure 4).

Of the resolved COVID-19 study participants collected, 11 (7.2%) were asymptomatic. Six of the 11 (54.5%) asymptomatic subjects did not produce any anti-S protein or anti-RBD antibodies (Figure 4). Six of the asymptomatic subjects were in the category of RT-PCR-positive/antibody-negative with a high Ct value.

**Figure 4:**
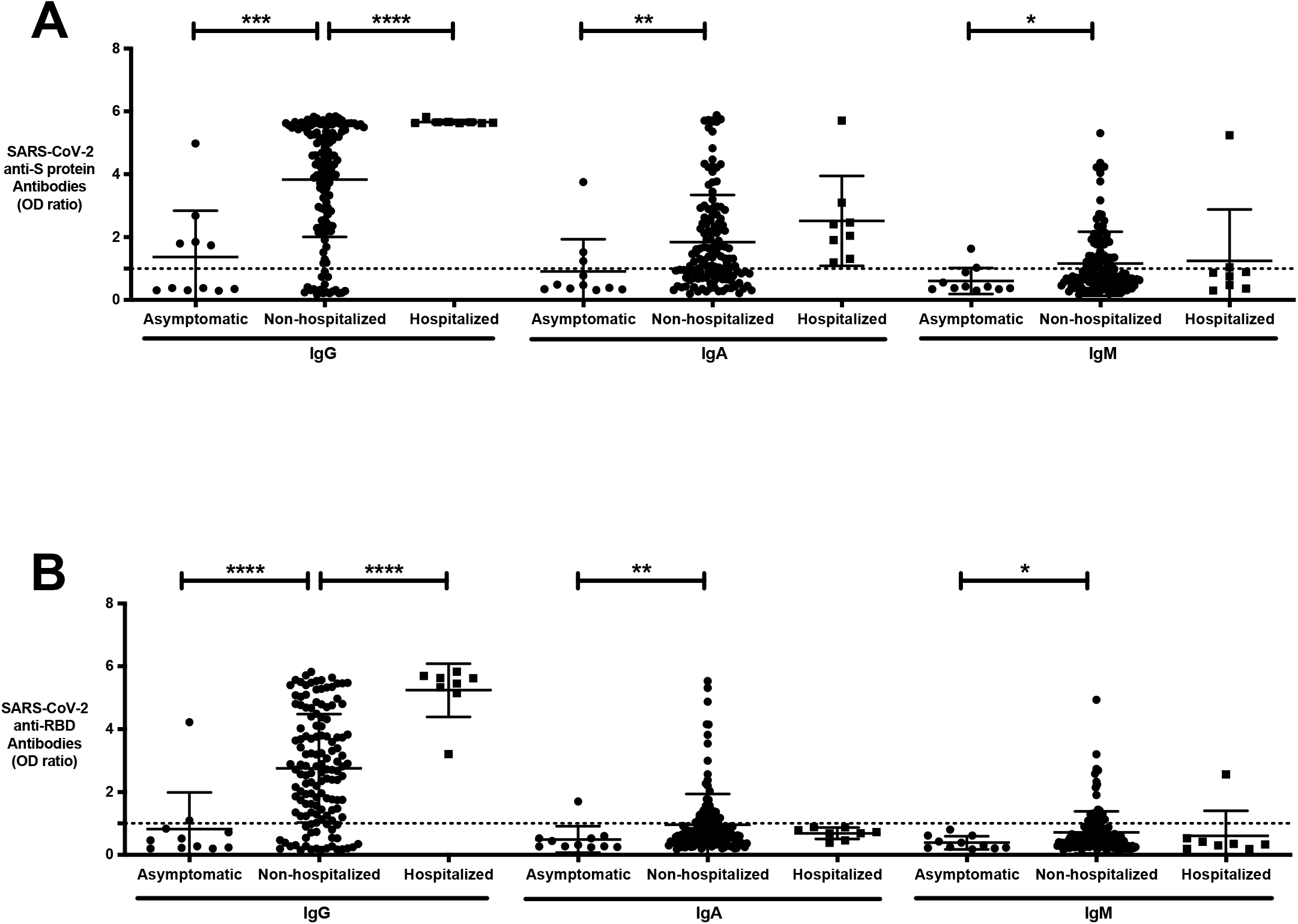
Comparing anti-SARS-CoV-2 IgG, IgA, and IgM responses to S protein and RBD antigens in asymptomatic, non-hospitalized and hospitalized resolved subjects. (A) Anti-S protein IgG, IgA, IgM and (B) anti-RBD IgG, IgA, and IgM of the asymptomatic (n=11), non-hospitalized resolved subjects (n=134) and hospitalized resolved subjects (n=8) were profiled using the SARS-CoV-2 ELISA. Values are shown as a ratio of observed optical density to the determined assay cut-off optical density. All hospitalized subjects had detectable anti-S protein IgG, anti-RBD IgG and anti-S protein IgA antibodies in their serum. The levels of anti-S protein and anti-RBD IgG (mean OD_405_ ratio) in resolved subjects who were hospitalized were significantly higher than the non-hospitalized resolved population. Values above 1 ratio are considered positive in the SARS-CoV-2 ELISA. * *p*<0.05, \*\**p*<0.01, \*\*\**p*<0.001, \*\*\*\**p*<0.0005.

## Discussion

Understanding the characteristics of the SARS-CoV-2 antibodies will inform on the seroprevalence in communities and portions of the COVID-19 immune response. We describe initial cross-sectional results of our longitudinal study of quantitative and functional SARS-CoV-2 antibodies in resolved subjects. The expression of IgG, IgA and IgM antibodies reactive to the immunogenic S protein and the RBD varied based on infection of SARS-CoV-2, severity of the disease, and time delay from onset of symptoms to blood draw. We also observed temporal and functional differences in anti-SARS-CoV-2 antibodies based on Ig classes and antigen.

Antibodies to SARS-CoV-2 were found in 131/153 (85.6%) subjects who tested positive in the RT-PCR. In 22 of 153 (14.4%) resolved subjects who tested RT-PCR-positive, no antibodies against SARS-CoV-2 were detected, which prompted further investigation using Ct values. Of 18 samples of whom Ct values were available, 17 had high Ct values (32.29 ± 4.647, range = 16.00 to 37.38, Figure 1D). The variation in antibody response in RT-PCR-positive subjects may be due to multiple contributing factors including the size of the viral inoculum, the genetic background of patients, and the presence of concomitant pathological conditions [27]. Studies have reported that after COVID-19 infection in some subjects, no antibodies can be detected in circulation either because they have waned quickly, or that their immune response is dependent on T cell responses [28-30]. Another factor is disease severity in the detection of SARS-CoV-2 antibodies. Strong neutralizing antibody responses may require more extensive affinity maturation, which is detected more in COVID-19 patients with severe disease symptoms [27, 31, 32]. Therefore, it is possible that the inability to detect antibodies in RT-PCR-positive subjects may be due to an infection insufficient of magnitude or durability [33]. Another possibility can be that the RT-PCR-positive/antibody-negative subjects had high Ct values at the limit of detection for the assay and did not actually contract the SARS-CoV-2 virus. One study showed there is reduced probability of cultivable viral cultures from samples with a Ct > 24 or when samples are obtained >8 days after symptom onset [34], and another study showed Ct values above 33-34 were not associated with cell culture viral recovery [35]. The developed ELISA for all Ig classes against S protein and RBD antigens has a sensitivity of 97.1% after removal of these 17 subjects with indeterminate test results from analysis.

Two subjects enrolled in this study were categorized as RT-PCR-negative based on a second RT-PCR test. Their initial RT-PCR test was positive, but retested RT-PCR-negative within 1-2 days after the first result due to the initial test having a high Ct value positive result without any symptoms or contact history. It is possible that some of the RT-PCR-positive/antibody-negative subjects with high Ct values, would have tested negative in a repeated RT-PCR test as well. A study showing repeat testing of the same subjects describes 6.8% of participants who initially test positive, tested negative in a follow-up RT-PCR test [36]. Repeating tests can reduce false-positive results, especially in those who have minimal indicators for having been infected. Another way to distinguish possible false-positive results in the RT-PCR is to use a combination of RT-PCR testing and antibody testing to improve the accuracy of COVID-19 diagnosis. One study utilizing rapid antigen diagnostic tests in combination with IgM/IgG detection, identified more subjects with COVID-19 admitted in an emergency department than when the assays are performed separately [37].

Conversely, SARS-CoV-2 antibodies were detected in three (5.5%) subjects in the cohort who tested RT-PCR-negative. Detection of SARS-CoV-2 antibodies in these samples may be a result of cross-reactivity of the antibodies with seasonal coronaviruses from previous infections [38, 39]. In the pre-COVID-19 samples (n=520), 11 (2.1%) had cross-reactivity to the SARS-CoV-2 antigens. Additionally, one of the three subjects who was RT-PCR-negative and positive for SARS-CoV-2 antibody was tested in the RT-PCR 113 days after initial symptom onset, which is later than the optimal timing since symptom onset for RT-PCR testing, possibly being the reason for a false-negative RT-PCR test. Taking into account the RT-PCR-negative cohort of subjects, we calculate the overall specificity of the in-house ELISA to be 96.7%.

Antibody kinetics reported previously in SARS-CoV-2 infected subjects suggest that titers of the virus-specific IgG and IgM antibodies increase 21 days post symptom onset [18, 40]. However, other studies indicate conflicting evidence on whether IgG and neutralizing antibody levels persist or begin to decline in a high proportion of recovered subjects within 2-3 months after infection [18]. Our study found that IgM positivity was lower than that of IgG after infection [18, 38, 41, 42]. Minimal differences were also observed in the percentage of anti-S protein and anti-RBD present in participants drawn at various times over a 3-month period. Furthermore, anti-SARS-CoV-2 antibodies could be found in the circulation of some resolved subjects 200 days post-symptom onset. Resolved subjects who were asymptomatic but RT-PCR-positive, had the lowest titre of SARS-CoV-2 antibodies.

Our study used whole live SARS-CoV-2 viruses in the neutralization assay which was able to determine the functional inhibitory capacity of antibodies against all antigens of SARS-CoV-2. Neutralizing antibody titres have recently been shown to wane fairly rapidly in some studies and levels were found to be variable in recovered subjects [19, 21, 43-45]. The U.S. Food and Drug Administration (FDA) recommends that the titer of neutralizing antibodies in convalescent plasma should be at least 1/160, but a 1/80 titer is acceptable in the absence of a better match for use in convalescent plasma therapy [46]. Most resolved COVID-19 subjects were found to have developed levels of SARS-CoV-2-specific neutralizing antibodies similar to other cross-sectional studies [19, 44, 45]. Neutralizing SARS-CoV-2 antibodies correlated best with a positive anti-RBD IgG (r = 0.5157) and anti-S protein IgG antibody (r = 0.6010) result. However, at this time we do not know what relevant thresholds of neutralizing antibodies confer protection from infection. Of interest, it has been shown that although Fc-dependent effector functions are required for optimal protection, these are often mediated by non-neutralizing antibodies [47]. Neutralization assays are performed with serum, and thus it is not possible to define the relative contribution of each antibody class to neutralizing activity.

This study has limitations. Recall bias of dates and symptoms by the study participants may affect the interpretation of timing of virus detection in relation to symptom onset. For asymptomatic cases, the time when infection was acquired is not known. In conclusion, our study suggests that there is a variable antibody response in resolved subjects and a variable reduction in antibody positivity over time. The negative antibody results found in the SARS-CoV-2 ELISA in RT-PCR-positive samples may suggest a varied immune response that warrants further studies. Although serologic tests cannot be used as the primary diagnostic test, they may be used to support diagnosis of COVID-19 for persons who are tested later, outside the optimal RT-PCR window. The resolved subjects collected for this study are part of a larger longitudinal study that will provide further insight on antibody prevalence over time. Further studies with this longitudinal cohort will be critical in characterizing the nature and kinetics of SARS-CoV-2 antibodies and their ability to confer long-term immunity.

## Data Availability

The authors confirm that the data supporting the findings of this study are available within the article.

## Acknowledgements

We thank Erjona Kruja for technical assistance. Funding support for this work was provided by grants from the Ontario Research Fund (ORF), COVID-19 Rapid Research Fund (#C-191-2426729-NAZY) and by the Canadian Institute of Health Research (CIHR)-COVID-19 Immunity Task Force (CITF; #VR2-173204) awarded to Dr. Ishac Nazy, and Academic Health Sciences Organization (HAHSO) grant awarded to Dr. Donald M. Arnold (#HAH-21-02). This work was also supported, in part, by a Weston Family Microbiome Initiative Grant and a Canadian Institute of Health Research (CIHR) COVID-19 Rapid Response grant to Dr. Matthew S. Miller. Dr. Miller was also supported, in part, by a CIHR New Investigator Award and an Ontario Early Researcher Award. Ali Zhang is supported by a PSI Research Trainee Fellowship and a CIHR Canada Graduate Scholarships – Doctoral Award. Hannah D. Stacey was supported in part by an Ontario Graduate Scholarship.

## Cell Lines

SARS-CoV-2/SB3-TYAGNC strain information: https://wwwnc.cdc.gov/eid/article/26/9/20-1495_article

## Authorship Contributions

AH carried out the described studies, analyzed data, and wrote the manuscript. DMA designed the research and helped write the manuscript. JWS and AZ carried out the described studies, analyzed data, and wrote the manuscript. JCM assisted with experimentation, provided technical assistance, and helped write the manuscript. VTC provided technical assistance. HDS and JCA provided technical assistance and materials. ZC, BJH, RC, NI, DMEB, MSM, and JGK designed the research and helped write the manuscript. IN designed the research, interpreted data and wrote the manuscript. All authors reviewed and approved the final version of the manuscript.

## Disclosure of Conflicts of Interest

The authors declare no competing financial interests.

## Notes

### Competing Interest Statement

The authors have declared no competing interest.

### Author Declarations

This study was approved by the Hamilton Integrated Research Ethics Board (HIREB) and informed written consent was obtained from all participants.

### Summary of Updates

Added cross-sectional study data. Increased number in populations.

