## Supplemental data for "Characteristics of anti-SARS-CoV-2 antibodies in recovered COVID-19 subjects"

**Supplemental Methods:**

*Measuring inter-assay variability and repeatability*

To determine inter-assay reproducibility, pre-COVID-19 controls (n=4) and COVID-19 positive (n=4) patient serum samples were tested in at least four separate assays using the SARS-CoV-2 ELISA described above. Results were reported as an optical density (405 nm with reference 490 nm) for each antigen and antibody isotype (IgG, IgA and IgM). Values are represented as a ratio of observed optical density to the determined assay cut-off optical density. Values above 1 ratio is considered positive in the SARS-CoV-2 ELISA.

*Evaluation of viral inactivation treatments on antibody testing in ELISA*

Patient serum was inactivated to eliminate residual virus in serum samples. A subset of recovered COVID-19 positive (n=10) and pre-COVID-19 controls (n=5) were used to compare two viral inactivation methods in human sera: heat-treatment and treatment with the detergent Triton X-100 [28-30]. For heat-treatment, patient sera (0.5 mL) were incubated with rotational shaking at 56°C for 30 minutes and centrifuged for 10 minutes at 14,000 x g, after which the supernatant was collected [29]. In parallel, duplicate samples were mixed with Triton X-100 to a final concentration of 1% (v/v) [30]. The non-treated and treated patient samples were tested in the SARS-CoV-2 ELISA as previously described. We however did not test whether any virus was actually present in serum samples and whether the inactivation method was effective against virus.

*Inhibition of IgG anti-RBD binding*

To confirm the specificity of antibodies detected by the ELISA, we inhibited the binding of selected serum samples with excess fluid-phase RBD. One antibody-positive pre-COVID-19 control and COVID-19 positive (n=10) patient serum samples were tested in the SARS-CoV-2 ELISA described above. Samples diluted to a working concentration (1/100) were incubated with 10 times molar excess of RBD or the equivalent volume of PBS for 1 hour at room temperature before testing for antibodies in the standard assay as described.

*Comparing the in-house SARS-CoV-2 ELISA to commercially available assays*

A subset of COVID-19 positive patient samples (n=9) and COVID-19 negative patient samples (n=5) was tested in the commercially available EUROIMMUN Anti-SARS-CoV-2 ELISA that measures anti-S1 IgG and IgA and the results were compared to our in-house assay. The same set of COVID-19 positive and pre-COVID-19 controls was also tested by the Hamilton Regional Laboratory Medicine Program (HRLMP) in the Ortho Clinical Diagnostics COVID-19 IgG Antibody Test that measures anti-S protein IgG for comparison.

*Production of recombinant SARS-CoV-2 S protein and RBD*

Plasmids encoding mammalian cell codon optimized sequences for SARS-CoV-2 full-length S protein and the RBD were generously gifted from the lab of Dr. Florian Krammer (Ichan School of Medicine at Mount Sinai, NY, NY, USA). In brief, proteins were produced in Expi293 cells (ThermoFisher, Waltham, MA, USA) using manufacturer instructions. Post-transfection, when culture viability dropped to 40%, supernatants were collected and centrifuged at 500 x g for 5 minutes to remove cell debris. The supernatant was then incubated with shaking overnight at 4°C with 1 mL of nickel-nitrilotriacetic acid (Ni-NTA) agarose resin (Qiagen, Hilden, Germany) per

25 mL of transfected cell supernatant. The following day, 10 mL polypropylene gravity flow columns (Qiagen) were used to elute the protein. S and RBD proteins were concentrated in Amicon centrifugal units (Millipore, Burlington, MA, USA), 10 kDa and 50 kDa respectively, prior to being resuspended in phosphate buffered saline (PBS). The purified proteins were then analyzed using SDS-PAGE.

*PCR Cycle count threshold (Ct) values of Resolved COVID19 samples*

Samples were analyzed from nasal or throat swabs and RT-PCR was measured at the Hamilton Regional Laboratory Medicine Program virology lab using an in-house RT-PCR targeted against the 5’ UTR and the E gene. The Ct values were determined by the number of cycles the PCR goes through before detection occurs. RNaseP was run on each assay to ensure specimen adequacy. The RT-PCR is a qualitative assay where the cut-offs change based on data collected over time and where the number of targets detected are also taken into consideration, along with examination of the amplification curve to determine positivity of a sample.

### Supplemental Figures:

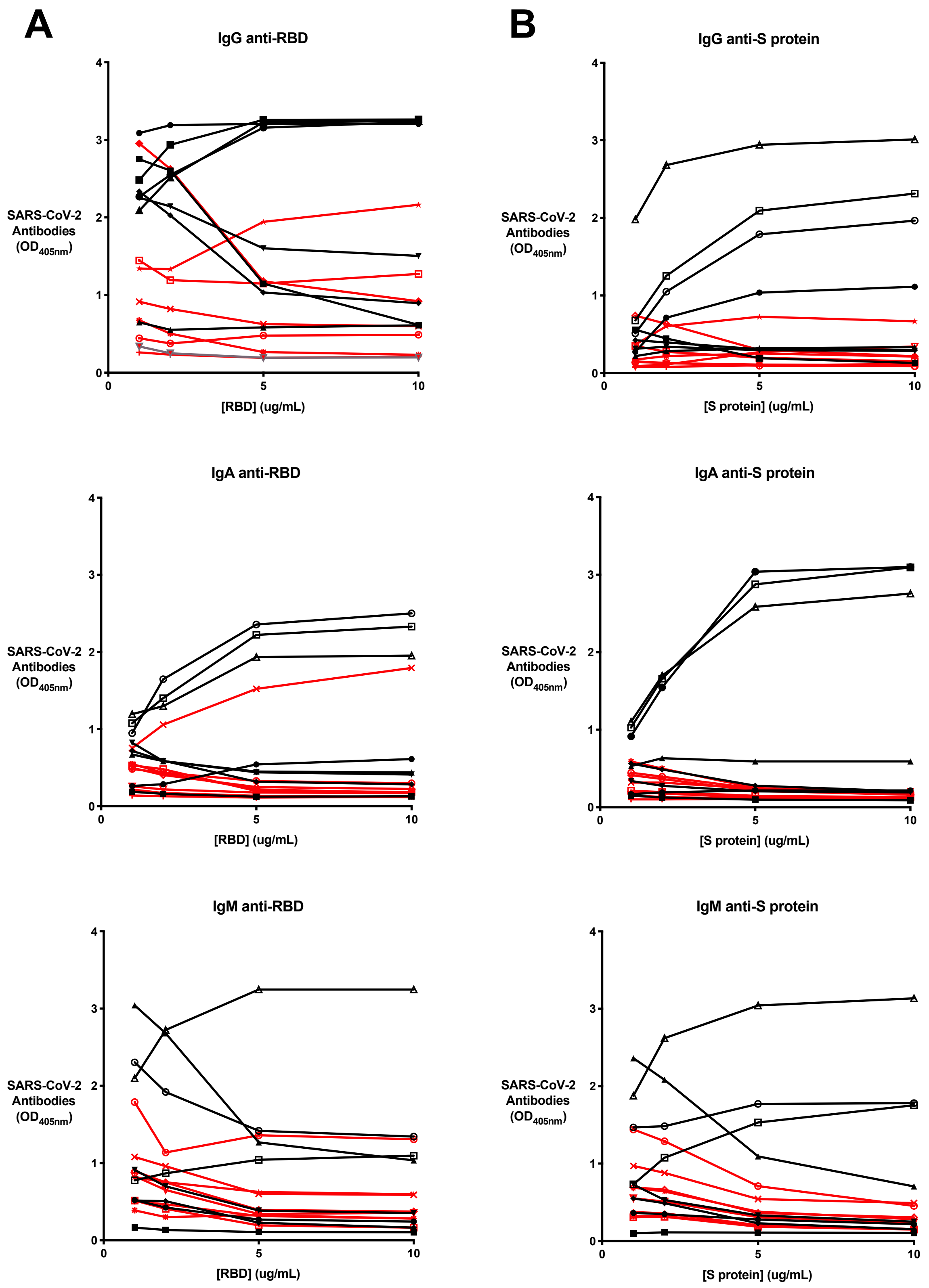

**Figure S1: Optimizing antigen concentration for the SARS-CoV-2 ELISA.** Binding of (A) anti-RBD and (B) anti-S protein IgG, A, and M was measured in the SARS-CoV-2 ELISA with plates coated at 1, 2, 5 or 10 μg/mL of the corresponding antigen. Black lines indicate recovered COVID-19 positive patient samples tested (n=8) and red lines represent pre-COVID-19 controls tested (n=8). RBD at 2 μg/mL and S protein at 5 μg/mL were the optimal concentrations used that yielded the greatest separation between OD values of COVID-19 positive and pre-COVID-19 controls results. The optimal serum dilution in 1% skim milk that provided the best separation between negative and positive results was 1/100.

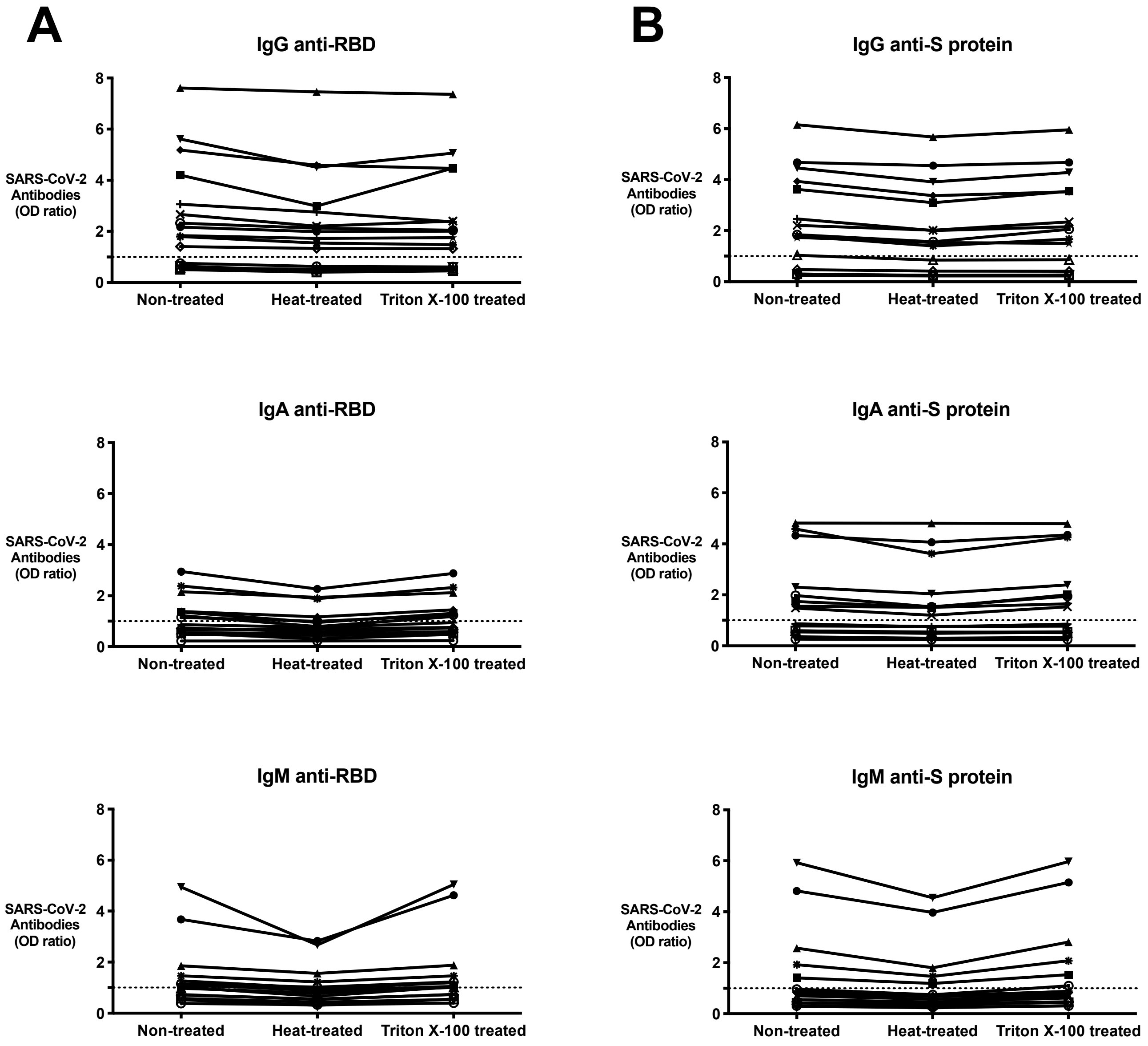


**Figure S2: Comparing the effect of heat-treatment and Triton X-100 treatment on assay performance.** Reactivity of (A) anti-RBD and (B) anti-S protein IgG, A, and M after heat-treatment or Triton X-100 treatment was measured using the SARS-CoV-2 ELISA. Serum samples used for this study included recovered COVID-19 positive patient samples (n=10) and pre-COVID-19 controls (n=5). Values are shown as a ratio of determined optical density to the determined assay cut-off optical density. Values above 1 are considered positive in the SARS-CoV-2 ELISA. The IgG signals were decreased in 12 of 15 samples (66.7%) by an average

13.54±6.73% after heat-treatment, but none of these samples decreased below the cut-off. Signals for IgA against RBD and S protein decreased in 12 of 15 samples (80.0%) by an average 13.85±9.55% after heat-treatment. With heat treatment, for IgM anti-RBD and IgM anti-S protein from COVID-19 positive patients and pre-COVID-19 controls, there was a reduction in OD by an average 21.0±8.06% after heat-treatment. Four of 10 COVID-19 positive samples IgM ODs decreased below the assay cut-off after heat-treatment. IgG signals in anti-RBD and anti-S protein were decreased in 14 of 15 samples by an average level of 8.03±7.59% after treatment with Triton X-100. ODs for IgA against RBD and S protein decreased in 7 of 15 samples by an average 4.14±3.38% and increased in the 8 other samples by an average level of 6.74±4.01% after treatment with Triton X-100. With Triton X-100 treatment, IgM anti-RBD and IgM anti-S protein had an average increase in OD by 6.4±5.64% in 11 of 15 samples and OD decreased by 3.11±2.68% in 4 of 15 samples. Treatment with Triton X-100 had minimal effect on the reactivity of the samples. None of the COVID-19 positive samples tested with Triton X-100 treatment decreased below the cut-off after treatment. None of the measured pre-COVID-19 controls became higher than the assay cut-off value after heat-inactivation or treatment with Triton X-100.

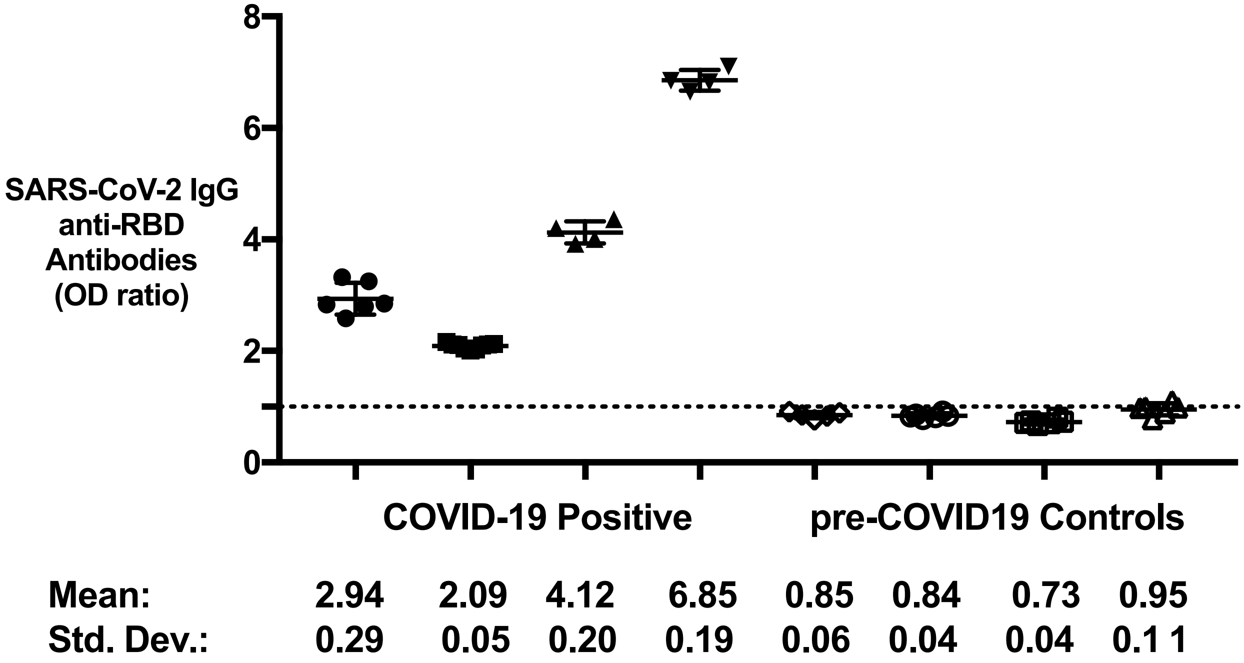


**Figure S3: Determining inter-assay variability of the SARS-CoV-2 ELISA.** Binding of anti-RBD IgG for a subset of COVID-19-positive (n=4) and pre-COVID-19 controls (n=4) were repeat tested at least 4 consecutive times to determine any variability between assays. Filled black symbols represent recovered COVID-19 positive patients and open symbols represent pre-COVID-19 controls. Values are represented as a mean optical density reading with standard deviation at 405 nm. The samples tested for variability using resulted in similar readings and low standard deviations for all repetitions. There is minimal inter-assay variability with repeat testing in anti-RBD IgG on average, results deviated by 4.86±3.53% in COVID-19 positive and 7.14±3.03% in pre-COVID-19 controls. This trend was similar for anti-RBD IgA (7.60±3.75% COVID-19 positive; 5.19±3.02% pre-COVID-19 controls) and IgM (4.21±2.64% COVID-19 positive; 8.58±4.63% pre-COVID-19 controls) and anti-S protein IgG (6.16±5.70% COVID-19 positive; 8.82±2.43% pre-COVID-19 controls), IgA (3.30±2.92% COVID-19 positive;

12.83±4.17% pre-COVID-19 controls), and IgM (6.08±2.68% COVID-19 positive; 12.37±4.18% pre-COVID-19 controls).

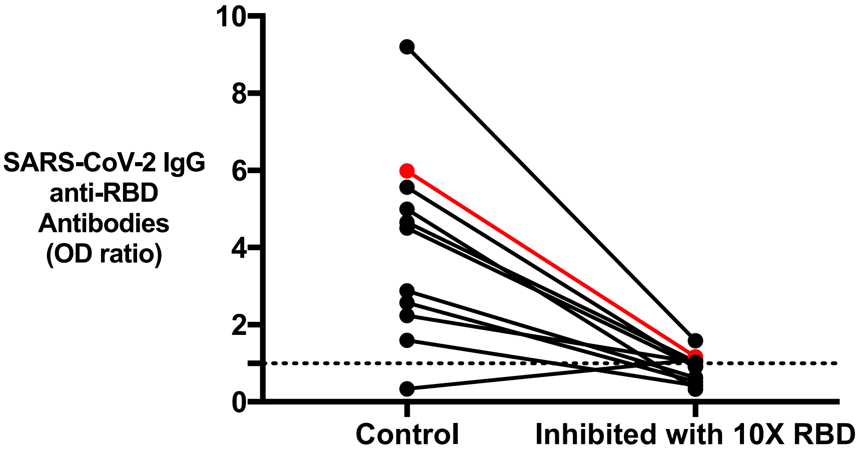


**Figure S4: Inhibition of IgG anti-RBD binding using excess RBD in solution.** Binding of IgG anti-RBD after inhibition with excess RBD in solution to show the specificity of antibodies to RBD antigen in the SARS-CoV-2 ELISA. Black lines indicate recovered COVID-19 patient samples tested (n=10) and red line represent one antibody-positive pre-COVID-19 control tested. The one pre-COVID-19 control tested positive for IgG anti-RBD antibodies that was also inhibited using excess RBD. All COVID-19 positive patient anti-RBD binding was inhibited on average 78.73±10.12% using excess RBD. One pre-COVID-19 control who tested positive for an anti-RBD IgG was also inhibited to a similar degree (80.51%) as COVID-19 positive samples with excess RBD in solution.

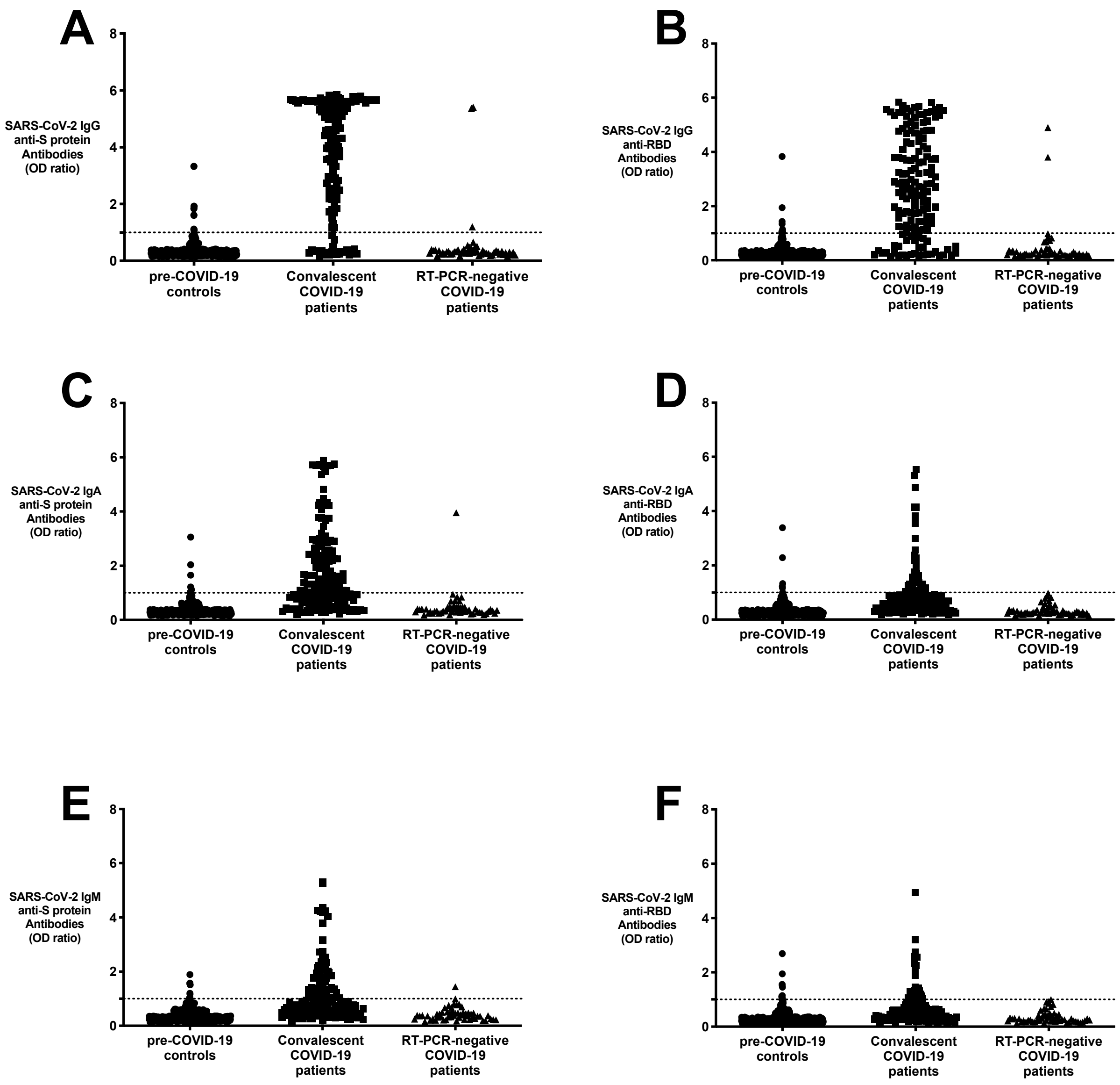


**Figure S5:** ***Cross-sectional analysis of IgG, IgA, and IgM responses to S-protein and RBD antigens of SARS-CoV-2 in serum.*** Anti-S protein (A) IgG, (C) IgA, (E) IgM and anti-RBD (B) IgG, (D) IgA, and (F) IgM of the pre-COVID-19 (n=520), resolved (n=156), and RT-PCR-negative (n=53) populations were profiled by the optimized SARS-CoV-2 ELISA. Values are shown as a ratio of observed optical density to the determined assay cut-off optical density. Values above 1 ratio are considered positive in the SARS-CoV-2 ELISA. Most pre-COVID-19 controls had only background reactivity for both the full-length S protein and RBD. Of the 153 resolved COVID-19 subjects tested, 131 tested positive for antibodies against SARS-CoV-2 (IgG, IgA, or IgM antibodies against the S protein or RBD, Table 2) and 22 did not have detectable anti-SARS-CoV-2 antibodies.

**Table S1: Comparing results from the in-house SARS-CoV-2 ELISA to two commercial assays**

| Sample ID | In-house SARS-CoV-2 ELISA (anti-S protein or anti-RBD IgG/A/M) | Ortho Clinical Diagnostics anti-S protein IgG | EUROIMMUN anti-S1 IgG or IgA |
|---|---|---|---|
| **C001** | Positive | Positive | Positive |
| **C003** | Negative | Negative | Negative |
| **C004** | Positive | Positive | Positive |
| **C005** | Positive | Positive | Positive |
| **C008** | Negative | Negative | Negative |
| **C009** | Negative | Negative | Positive (IgA) |
| **C010** | Negative | Negative | Borderline Positive (IgG) |
| **C019** | Negative | Negative | Negative |
| **C021** | Positive | Negative | Negative |
| **C023** | Negative | Negative | Negative |
| **C031** | Positive | Positive | Positive |
| **C039** | Positive | Positive | Positive |
| **C041** | Positive | Positive | Positive |
| **C050** | Positive | Positive | Positive |

Samples highlighted were found to have different results between the in-house SARS-CoV-2 ELISA, the Ortho assay, and the EUROIMMUN assay. The EUROIMMUN Anti-SARS-CoV-2 ELISA measures IgG or IgA antibodies to the S1 protein. The Ortho Clinical Diagnostics

COVID-19 IgG Antibody Test measures IgG antibodies to the S protein.  We found that the in-house SARS-CoV-2 ELISA agreed with the Ortho assay performed by the clinical laboratory (HLRMP), in 14/14 samples (Table 2) for IgG antibodies to the S protein. In one COVID-negative patient sample, the in-house assay detected weak IgG anti-RBD antibodies only, and no other reactivity, which correlated with the Ortho test. The EUROIMMUN assay detected SARS-CoV-2 antibodies in two COVID-19 positive patient samples where the in-house ELISA and Ortho assay did not, one of which was borderline positive for IgG anti-S1.
